# Clinical Characteristics and Durations of Hospitalized Patients With COVID-19 in Beijing: A Retrospective Cohort Study

**DOI:** 10.1101/2020.03.13.20035436

**Authors:** Wen Zhao, Xiangyi Zha, Ning Wang, Dongzeng Li, Aixin Li, Shikai Yu

## Abstract

**Objective:** To give the information on clinical characteristics and different durations of COVID-19 and to identify the potential risk factors for longer hospitalization duration.

**Methods:** In this retrospective study, we enrolled 77 patients (mean age: 52±20 years; 44.2% males) with laboratory-confirmed COVID-19 admitted to Beijing YouAn Hospital during 21^st^ Jan and 8^th^ February 2020. Epidemiological, clinical, and radiological data on admission were collected; complications and outcomes were followed up until 26^th^ February 2020. The study’s endpoint was the discharge within two weeks. Cox proportional-hazards regression was performed to identify risk factors for longer hospitalization duration.

**Results:** Of 77 patients, there were 34 (44.2%) males, 24 (31.2%) with comorbidities, 22 (28.6%) lymphopenia, 20 (26.0%) categorized as severe patients, and 28 (36.4%) occurred complications. By the end of follow-up, 64 (83.1%) patients were discharged home, 8 remained in hospital and 5 died. 36 (46.8%) patients were discharged within 14 days and thus reached the study endpoint, including 34 (59.6%) of 57 non-severe patients and 2 (10%) of 20 severe patients. The overall cumulative probability of the endpoint was 48.3%. Hospital length of stay and duration of exposure to discharge for 64 discharged patients were 13 (10-16.5) and 23 (18-24.5) days, respectively. Multivariable stepwise Cox regression model showed that bilateral pneumonia on CT scan, shorter time from the illness onset to admission, severity of disease and lymphopenia were independently associated with longer duration of hospitalization.

**Conclusions:** COVID-19 has significantly shorter duration of disease and hospital length of stay than SARS. Bilateral pneumonia on CT scan, shorter period of illness onset to admission, lymphopenia, severity of disease are the risk factors for longer hospitalization duration of COVID-19.

**Significance Statement:** In this study, we reported that the average hospital length of stay for discharged patients with COVID-19 is 13 days and the average time of clinical course of COVID-19 is 23 days, both of which are significantly shorter than that of SARS. The risk factors for longer hospitalization duration of COVID-19 include bilateral pneumonia on CT scan, shorter period of illness onset to admission, lymphopenia, and severity of disease. There findings might be helpful for the countries or territories facing the threat of COVID-19 to well prepare and rebalance their medical resources.

## Introduction

Since started in the last month of 2019, the outbreak of the SARS-Cov-2 infection has successfully attracted attention of the whole world. By virtue of its highly contagious attribution, this novel coronavirus has spread across over 113 countries/territories and caused 118,319 infected cases with 4292 deaths, as of 11^th^ March 2020 and the numbers are still rapidly increasing^[1]^. Moreover, the threat of SARS-Cov-2 to global health is rapidly increasing, which made the World Health Organization declare COVID-19 a ‘Pandemic’. Undoubtedly, preparedness for the virus strike is necessary and critically important, especially for those countries at high risk but with relatively weaker public health systems. Under this circumstance, the experience of containment from peer countries and territories and comprehensive research on COVID-19 would undoubtedly help. Many previous studies have generally reported the epidemiological features, clinical characteristics and virology of SARS-Cov-2 ^[2–8]^. But the features and characteristic of the disease may present differently among different locations and over time^[9]^, since the pathogen, SARS-Cov-2, may evolve after transmissions ^[10]^. It thus is important to closely follow these changes for better containment and management of COVID-19. In addition, the durations of COVID-19 (e.g., the average time from exposure to recovery and hospitalization duration) which is important for understanding this disease and useful for the preparedness and response correctly to COVID-19, were not reported, mainly because most patients included in the previous studies still remained in hospital as the study being done. Here, we presented the clinical data of 77 hospitalized patients with COVID-2019 in YouAn Hospital (Beijing, China) with the aim of giving the information about clinical characteristics and different durations of COVID-19: incubation period, illness onset to first hospital, illness onset to discharge, exposure to discharge, and hospitalization duration. Additionally, this paper also aims to identify the potential risk factors for longer hospitalization duration of COVID-19.

## Methods

### Data sources and collection

We collected and retrospectively reviewed the medical records and compiled data of all 77 hospitalized patients with laboratory-confirmed COVID-19 in Beijing YouAn Hospital, Beijing, China between 21^st^ Jan and 8^th^ Feb 2020. COVID-19 was diagnosed in accordance with the interim guidance from the World Health Organization ^[11]^. A confirmed case of COVID-19 was defined as a positive result on real-time reverse-transcriptase-polymerase-chain-reaction (RT-PCR) assay of Nasopharyngeal swab samples. The local ethic review board of the hospital approved this study and waived informed consent for each patient, considering the retrospective nature of this study.

Epidemiological, clinical, laboratory and radiographic data and outcome data were extracted from electronic medical records. In detail, the following information was collected: demographics, exposure and medical history, coexisting diseases, symptoms and signs, laboratory and CT scan findings on admission, complications, and outcome data during follow-up. Complications and clinical outcome were followed up until 26 Feb 2020. Each case was categorized as non-severe or severe at the time of admission and acute respiratory distress syndrome (ARDS) was defined according to the guidance of WHO for COVID-19 ^[11]^ Acute kidney injury according to the Kidney Disease: Improving Global Outcomes definition.^[12]^ The durations of illness onset to first hospital admission and to discharge were calculated, as well as hospitalized duration (hospital length of stay). The durations of exposure to illness onset and to discharge were calculated in those who had a single point of exposure history and could provide specific date.

### Study outcomes

It is currently unclear about the normal length of hospital stay for patients with COVID-19 discharged home alive. Retrospectively, the average hospital length of stay of 64 discharged patients in our study is 13 days. Thus, we set the discharge within two weeks (14 days) as the endpoint of our study and accordingly divided the study patients into two groups: with endpoint (shorter hospitalization duration) and without endpoint (longer hospitalization duration). Fitness for discharge was based on abatement of fever for at least three days, with improved evidence on chest radiography and at least 2 consecutive negative PCR assay results with more than 24 hours apart ^[13]^.

### Statistical analysis

Continuous variables were expressed as means ± standard deviations (SD) for the normally distributed data or median with interquartile (IQR) for the skewed data. Correspondingly, two-sample independent t-test and Mann-Whitney U test were used to detect the difference between groups. Categorical variables were described as number (%) and compared by χ^2^ test or Fisher’s exact test as appropriate. Kaplan-Meier method was used to estimate the cumulative probability of the endpoint. Cox proportional-hazards regression model was conducted to determine the potential risk factors associated with the endpoint. Statistical significance was defined as P<0.05. All analyses were done with SAS software, version 9.4.

## Results

### Epidemiological and clinical characteristics

The epidemiological and clinical characteristics of the study patients are shown in Table 1. The patients in this study had a very wide age span (1-94 years) with mean value at 52 years and there were 34 (44.2%) males. A history of exposure to Wuhan was found in most patients (70.1%), especially in the initial stage of the COVID-19 spreading in Beijing, as shown in Figure 1. No patients had histories of exposure to the Huanan market and/or wildlife animals. There were 33 patients (42.9%) involved in family clusters of infections and 2 healthcare workers (2.6%). Fever was the most common symptom at the onset of illness, followed by cough and fatigue; digestive symptoms such as diarrhea were rare. Two patients were asymptomatic on admission. Twenty-four patients (31.2%) presented coexisting disorders, including hypertension (20.8%), cardiovascular disease (11.7%), diabetes (7.8%), among others.

**Table 1.** Characteristics of 77 patients with COVID-19 in Beijing, China.

| Variables | All patients<br>(n=77) | Severity of disease |  | P | Presence of endpoint |  | P |
| --- | --- | --- | --- | --- | --- | --- | --- |
|  |  | Non-severe<br>(n=57, 74.0%) | Severe<br>(N=20, 26.0%) |  | Yes<br>(n=36, 46.8%) | No<br>(n=41, 53.2%) |  |
| Age, years | 52±20 | 45±17 | 69±15 | <0.001 | 43±16 | 59±20 | <0.001 |
| Age group, n (%) |  |  |  | <0.001 |  |  | 0.008 |
| 0-17 | 4 (5.2) | 4 (7.0) | 0 (0) |  | 3 (8.3) | 1 (2.4) |  |
| 18-39 | 21 (27.3) | 20 (35.1) | 1 (5.0) |  | 15 (41.7) | 6 (14.6) |  |
| 40-64 | 30 (39.0) | 25 (43.9) | 5 (25.0) |  | 14 (38.9) | 16 (39.0) |  |
| 65-79 | 15 (19.5) | 6 (10.5) | 9 (45.0) |  | 3 (8.3) | 12 (29.3) |  |
| ≥80 | 7 (9.1) | 2 (3.5) | 5 (25.0) |  | 1 (2.8) | 6 (14.6) |  |
| Sex, n (%) |  |  |  | 0.26 |  |  | 0.38 |
| Male | 34 (44.2) | 23 (40.4) | 11 (55.0) |  | 14 (38.9) | 20 (48.8) |  |
| Female | 43 (55.8) | 34 (59.7) | 9 (45.0) |  | 22 (61.1) | 21 (51.2) |  |
| Exposure history, n (%) |  |  |  | 0.07 |  |  | 0.06 |
| Wuhan exposure | 54 (70.1) | 44 (77.2) | 10 (50.0) |  | 28 (77.8) | 26 (63.4) |  |
| Contact with other suspected or confirmed cases | 15 (19.5) | 8 (14.0) | 7 (35.0) |  | 3 (8.3) | 12 (29.3) |  |
| No known exposure | 8 (10.4) | 5 (8.8) | 3 (15.0) |  | 5 (13.9) | 3 (7.3) |  |
| Family cluster, n (%) | 33 (42.9) | 24 (42.1) | 9 (45.0) | 1.00 | 12 (33.3) | 21 (51.2) | 0.14 |
| Health worker, n (%) | 2 (2.6) | 2 (3.5) | 0 (0) | 1.00 | 1 (2.8) | 1 (2.4) | 1.00 |
| Symptoms on admission |  |  |  |  |  |  |  |
| Fever | 66 (85.7) | 47 (82.5) | 19 (95.0) | 0.27 | 29 (80.6) | 37 (90.2) | 0.23 |
| Productive cough | 25 (32.5) | 16 (28.1) | 9 (45.0) | 0.18 | 10 (27.8) | 15 (36.6) | 0.41 |
| Non-productive cough | 24 (31.2) | 17 (29.8) | 7 (35.0) | 0.78 | 12 (33.3) | 12 (29.3) | 0.78 |
| Fatigue | 21 (27.3) | 19 (33.3) | 2 (10.0) | 0.08 | 12 (33.3) | 9 (22.0) | 0.26 |
| Headache or dizziness | 10 (13.0) | 10 (17.5) | 0 (0) | 0.06 | 6 (16.7) | 4 (9.8) | 0.50 |
| Myalgia or arthralgia | 9 (11.7) | 9 (15.8) | 0 (0) | 0.10 | 4 (11.1) | 5 (12.7) | 1.00 |
| Chest tightness | 9 (11.7) | 5 (8.8) | 4 (20.0) | 0.23 | 4 (11.1) | 5 (12.2) | 1.00 |
| Shortness of breath | 16 (20.8) | 8 (14.0) | 8 (40.0) | 0.02 | 5 (13.9) | 11 (26.8) | 0.26 |
| Anorexia | 7 (9.1) | 4 (7.0) | 3 (15.0) | 0.29 | 3 (8.3) | 4 (9.8) | 1.00 |
| Nausea or vomiting | 6 (7.8) | 3 (5.3) | 3 (15.0) | 0.18 | 2 (5.6) | 4 (9.8) | 0.68 |
| Sore throat | 5 (6.5) | 5 (8.8) | 0 (0) | 0.32 | 2 (5.6) | 3 (7.3) | 1.00 |
| Nasal congestion or rhinorrhea | 8 (10.4) | 6 (10.5) | 2 (10.0) | 1.00 | 3 (8.3) | 5 (12.2) | 0.72 |
| Diarrhea | 1 (1.3) | 1 (1.8) | 0 (0) | 1.00 | 1 (2.8) | 0 (0) | 0.48 |
| asymptomatic | 2 (2.6) | 2 (3.5) | 0 (0) | 1.00 | 1 (2.8) | 1 (2.4) | 1.00 |
| Coexisting conditions, n (%) |  |  |  |  |  |  |  |
| Any | 24 (31.2) | 14 (24.6) | 10 (50.0) | 0.03 | 6 (16.7) | 18 (43.9) | 0.01 |
| Hypertension | 16 (20.8) | 8 (14.0) | 8 (40.0) | 0.02 | 3 (8.3) | 13 (31.7) | 0.01 |
| Diabetes | 6 (7.8) | 4 (7.0) | 2 (10.0) | 0.65 | 2 (5.6) | 4 (9.8) | 0.68 |
| Cardiovascular disease | 9 (11.7) | 3 (5.3) | 6 (30.0) | 0.008 | 1 (2.8) | 8 (19.5) | 0.03 |
| Cerebrovascular disease | 2 (2.6) | 1 (1.8) | 1 (5.0) | 0.45 | 0 (0) | 2 (4.9) | 0.50 |
| Respiratory disease | 6 (7.8) | 3 (5.3) | 3 (15.0) | 0.18 | 0 (0) | 6 (14.6) | 0.03 |
| Digestive disease | 8 (10.4) | 4 (7.0) | 4 (20.0) | 0.19 | 2 (5.6) | 6 (14.6) | 0.27 |
| Chronic renal disease | 5 (6.5) | 2 (3.5) | 3 (15) | 0.11 | 0 (0) | 5 (12.2) | 0.06 |
| Malignancy | 4 (5.2) | 3 (5.3) | 1 (5.0) | 1.00 | 1 (2.8) | 3 (7.3) | 0.62 |
| Severity of disease on admission |  |  |  |  |  |  | <0.001 |
| Non-severe | 57 (74.0) | / | / | / | 34 (94.4) | 23 (56.1) |  |
| Severe | 20 (26.0) | / | / | / | 2 (5.6) | 18 (43.9) |  |
| Complications during follow-up, n (%) |  |  |  |  |  |  |  |
| Any | 28 (36.4) | 19 (33.3) | 9 (45.0) | 0.42 | 9 (25.0) | 19 (46.3) | 0.06 |
| ARDS | 3 (3.9) | 0 (0) | 3 (15.0) | 0.02 | 0 (0) | 3 (7.3) | 0.24 |
| Shock | 1 (0) | 0 (0) | 1 (5.0) | 0.26 | 0 (0) | 1 (2.4) | 1.00 |
| Acute heart failure | 2 (2.6) | 0 (0) | 2 (10.0) | 0.06 | 0 (0) | 2 (4.9) | 0.50 |
| Acute kidney injury | 2 (2.6) | 1 (1.8) | 1 (5.0) | 0.45 | 1 (2.8) | 1 (2.4) | 1.00 |
| Liver dysfunction | 25 (32.5) | 19 (33.3) | 6 (30.0) | 0.78 | 9 (25.0) | 16 (39.0) | 0.19 |
| MODS | 1 (1.3) | 0 (0) | 1 (5.0) | 0.26 | 0 (0) | 1 (2.4) | 1.00 |
| Secondary infection | 3 (3.9) | 0 (0) | 3 (15.0) | 0.02 | 0 (0) | 3 (7.3) | 0.24 |

Continuous variables presented as Mean±SD if normally distributed, otherwise, as median (IQR). Categorical variables presented as numbers (percentages). ARDS, acute respiratory distress syndrome; MODS, multiple organ dysfunction syndrome.

**Figure 1.**
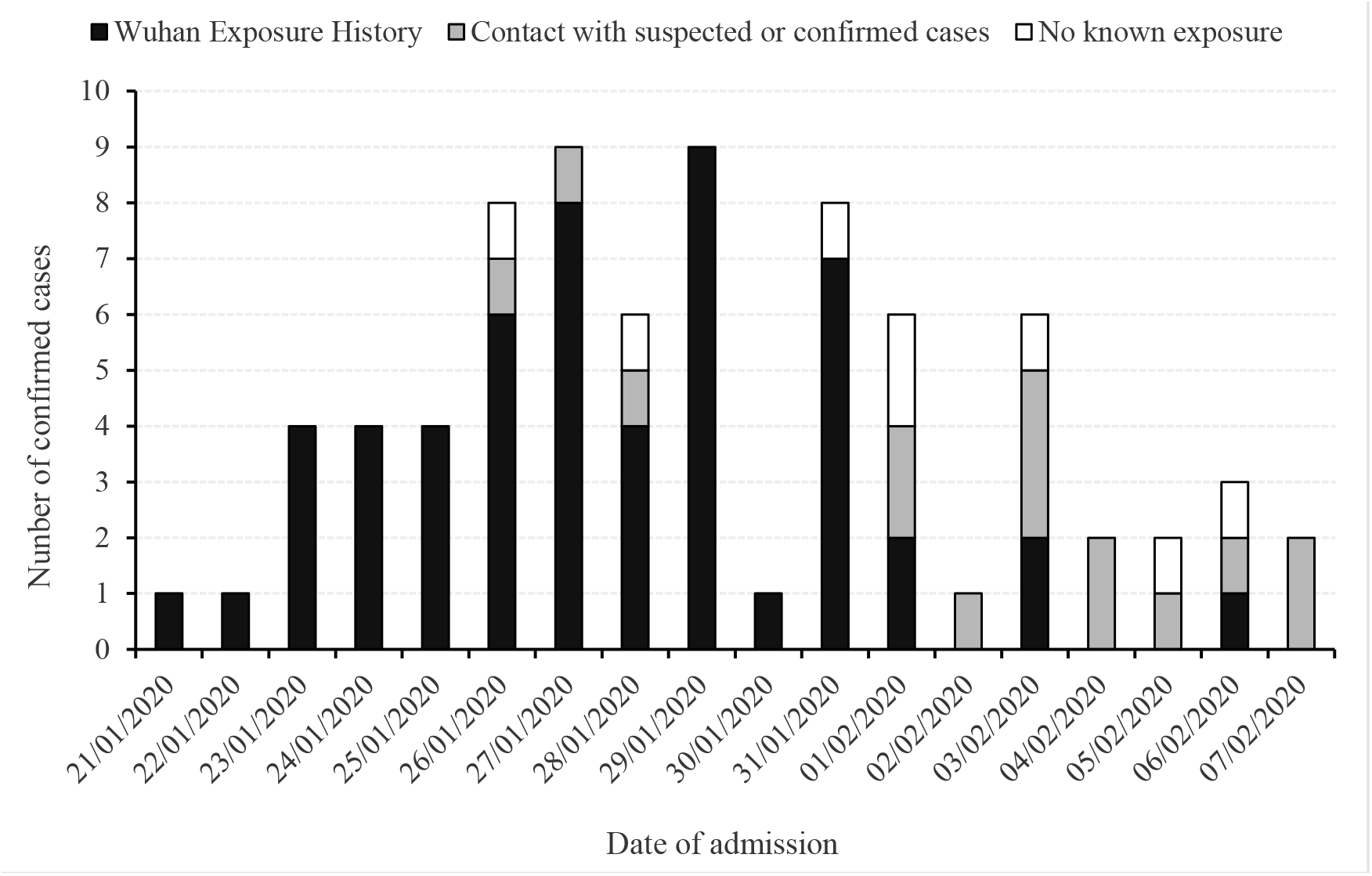
Date of admission and exposure histories of the 77 study patients with COVID-19.

There were 57 (74.0%) and 20 (26.0%) patients categorized as non-severe and severe patients, respectively. 28 patients (36.4) developed at least one complication, with liver dysfunction (32.5%) being the most common one and other complications were rare. Compared with non-severe patients, severe patients were older in age, more likely to present with shortness of breath in symptoms and have higher occurrence of hypertension and cardiovascular disease in terms of coexisting diseases.

### Laboratory and radiographic findings

The laboratory and radiographic findings on admission are presented in Table 2. White blood cell count, neutrophil and lymphocyte proportions were reduced in 35 (45.5%), 18 (23.4%) and 22 (28.6%) patients, respectively. Elevations of the count/proportion of these blood cells could also be observed in some patients. Significant increase of C-reactive protein was observed in most patients (64.9%). Of 73 patients with chest CT scans at the time of admission, 71 (97.3%) revealed abnormal results including 3 (4.1%) unilateral pneumonia and 68 (93.2%) bilateral pneumonia. In contrast to non-severe patients, severe patients showed significantly higher levels of white blood cell and neutrophil, but lower levels of lymphocyte, monocyte, C-reactive protein, lactate concentration, procalcitonin, creatine kinase-MB, myoglobin, and hypersensitive troponin I.

**Table 2.** Laboratory and radiographic findings.

| Variables | All patients<br>(n=77) | Severity of disease |  |  | Presence of endpoint |  | P |
| --- | --- | --- | --- | --- | --- | --- | --- |
|  |  | Non-severe patients<br>(n=58,74.0%) | Severe patients<br>(n=20,26.0%) |  | Yes<br>(n=36,46.8%) | No<br>(n=41,53.2%) |  |
| WBC count, $\times 10^9/L$ or n (%) | 4.04 (3.48-5.58) | 3.95 (3.45-4.64) | 5.92 (3.68-7.59) | 0.02 | 4.04 (3.51-4.91) | 4.04 (3.42-6.49) | 0.50 |
| <4 | 35 (45.5) | 29 (50.0) | 6 (31.6) | 0.02 | 17 (47.2) | 18 (43.9) | 0.40 |
| >10 | 2 (2.6) | 0 (0) | 2 (10.5) |  | 0 (0) | 2 (4.9) |  |
| Neutrophil, % or n (%) | 61.5±14.9 | 56.9±12.6 | 74.5±13.2 | <0.001 | 56.7±9.9 | 65.6±17.2 | 0.006 |
| <50 | 18 (23.4) | 17 (29.3) | 1 (5.3) | <0.001 | 11 (30.6) | 7 (17.1) | 0.003 |
| >70 | 23 (29.9) | 10 (17.2) | 13 (68.4) |  | 4 (11.1) | 19 (46.3) |  |
| Lymphocyte, % or n (%) | 27.5±13.0 | 31.2±11.7 | 16.9±10.6 | <0.001 | 31.5±9.1 | 24.0±14.9 | 0.008 |
| <20 | 22 (28.6) | 10 (17.2) | 12 (63.2) | <0.001 | 3 (8.3) | 19 (46.3) | 0.001 |
| >40 | 12 (15.6) | 11 (19.0) | 1 (5.3) |  | 7 (19.4) | 5 (12.2) |  |
| Monocyte, % or n (%) | 6.2 (4.8-8.0) | 6.7 (5.0-8.7) | 5.5 (3.9-6.7) | 0.01 | 6.75 (5.35-8.60) | 5.8 (4.5-7.8) | 0.10 |
| <3 | 4 (5.2) | 1 (1.7) | 3 (15.8) | 0.02 | 0 (0) | 4 (9.8) | 0.13 |
| >8 | 20 (26.0) | 18 (31.0) | 2 (10.5) |  | 11 (30.6) | 9 (22.0) |  |
| RBC count, $\times 10^{12}/L$ | 4.48±0.53 | 4.48±0.49 | 4.46±0.64 | 0.85 | 4.55±0.45 | 4.41±0.58 | 0.25 |
| Platelet count, $\times 10^9/L$ or n (%) | 190 (149-245) | 189 (152-245) | 198 (134-272) | 0.83 | 185 (156-248) | 192 (141-240) | 0.52 |
| <100 | 5 (6.5) | 2 (3.5) | 3 (15.0) | 0.11 | 2 (5.6) | 4 (7.3) | 0.75 |
| Haemoglobin, g/L | 137 (125-145) | 139 (127-144) | 137 (119-148) | 0.71 | 140 (126-146) | 136 (123-142) | 0.56 |
| CRP, mg/L or n (%) | 17.0 (4.6-51.1) | 13.0 (2.9-25.6) | 55.0 (16.5-110.0) | <0.001 | 16.6 (3.3-24.9) | 19.3 (6.3-75.1) | 0.08 |
| $\geq 10$ | 50 (64.9) | 32 (56.1) | 18 (90.0) | 0.006 | 23 (63.9) | 27 (65.9) | 0.86 |
| Prothrombin time, s | 12.7 (12.2-13.1) | 12.7 (12.2-13.1) | 12.7 (12.0-13.3) | 0.71 | 12.7 (12.2-13.0) | 12.7 (12.0-13.2) | 0.84 |
| APTT, s | 32.8 (30.4-34.7) | 33.1 (30.8-34.9) | 32.0 (28.9-34.7) | 0.28 | 33.0 (30.8-34.7) | 32.6 (29.8-34.7) | 0.59 |
| Po2, mmHg | 83.9 (72.9-107.0) | 82.5 (74.7-101.7) | 88.9 (64.6-110.0) | 0.54 | 82.2 (72.9-98.8) | 86.7 (71.1-115.1) | 0.70 |
| Pco2, mmHg | 31.1 (27.0-33.7) | 31.7 (28.5-34.4) | 28.3 (25.6-33.5) | 0.09 | 32.0 (27.6-36.8) | 29.8 (26.7-33.7) | 0.41 |
| Lactate concentration, mmol/L | 1.18 (0.89-1.68) | 1.07 (0.87-1.37) | 1.75 (1.16-1.96) | 0.002 | 1.05 (0.89-1.36) | 1.22 (0.94-1.79) | 0.19 |
| AST, U/L or n (%) | 29 (21-42) | 28 (21-38) | 36 (23-71) | 0.08 | 28 (19-34) | 34 (23-51) | 0.04 |
| >40 | 20 (26.0) | 11 (19.3) | 9 (45.0) | 0.04 | 4 (11.1) | 16 (39.0) | 0.005 |
| ALT, U/L or n (%) | 28 (20-46) | 28 (20-45) | 32 (19-58) | 0.37 | 26 (20-43) | 29 (20-51) | 0.28 |
| >40 | 26 (33.8) | 17 (29.8) | 9 (45.0) | 0.17 | 9 (25.0) | 17 (41.5) | 0.13 |
| Creatinine, $\mu\text{mol/L}$ or n (%) | 64 (54-78) | 63 (54-74) | 68 (52-84) | 0.79 | 62 (54-74) | 70 (54-82) | 0.38 |
| >133 | 2 (2.6) | 2 (3.5) | 0 (0) | 1.00 | 1 (2.8) | 1 (2.4) | 1.00 |
| Procalcitonin $\text{ug/L}$ or n (%) | 0.12 (0.10-0.14) | 0.11 (0.10-0.13) | 0.13 (0.11-0.17) | 0.03 | 0.11 (0.10-0.14) | 0.13 (0.11-0.14) | 0.01 |
| >0.5 | 1 (1.3) | 0 (0) | 1 (5.0) | 0.26 | 0 (0) | 1 (2.4) | 1.00 |
| Creatinine kinase, U/L | 80 (46-128) | 67 (45-118) | 115 (48-176) | 0.24 | 59 (44-95) | 101 (46-184) | 0.02 |
| Creatine kinase–MB, U/L | 0.33 (0.14-0.93) | 0.28 (0.09-0.47) | 0.87 (0.29-2.30) | 0.003 | 0.24 (0.07-0.40) | 0.54 (0.24-2.24) | <0.001 |
| Myoglobin, ng/mL | 41 (29-75) | 36 (28-57) | 74 (46-193) | 0.002 | 34 (28-51) | 57 (36-130) | 0.001 |
| hTnI, ng/mL or n (%) | 0.036±0.093 | 0.010 (0.010-0.010) | 0.021 (0.010-0.077) | <0.001 | 0.010±0.007 | 0.059±0.123 | 0.02 |
| >0.05 | 9 (11.7) | 2 (3.5) | 7 (35.0) | <0.001 | 0 (0) | 9 (22.0) | 0.003 |
| Chest CT findings (n=73), n (%) |  |  |  | 0.39 |  |  | 0.05 |
| Normal | 2 (2.7) | 2 (3.7) | 0 (0) |  | 2 (5.9) | 0 (0) |  |
| Unilateral pneumonia | 3 (4.1) | 3 (5.6) | 0 (0) |  | 3 (8.8) | 0 (0) |  |
| Bilateral pneumonia | 68 (93.2) | 49 (90.7) | 19 (100%) |  | 29 (85.3) | 39 (100) |  |

Continuous variables presented as Mean ± SD if normally distributed, otherwise, as median (IQR). Categorical variables presented as numbers (percentages). WBC, white blood cell; RBC, red blood cell; CRP, C-reactive protein; APTT, Activated partial thromboplastin time; AST, aspartate aminotransferase; ALT, alanine aminotransferase; hTnI, hypersensitive troponin I.

### Durations of hospitalized patient with COVID-19

Of the 77 patients, twenty-three (29.9%) had a single episode of exposure history and could provide the specific date. As shown in Table 3, the median and interquartile range of incubation period for those patients was 4 (3-7) days. For all patients, time of illness onset to admission and hospital duration were 5 (3-6) and 13 (10-18) days, respectively. In terms of discharged patients, times of illness onset to discharge and of exposure point to discharge (available in 21 patients) were 18.5 (15-22) and 23 (17-25) days, respectively. In contrast to severe patients, non-severe patents showed shorter hospitalization duration (12 [10-16] vs. 18.5 [15-21] days), times of illness onset to discharge (18 [15-21] vs. 24.5 [22-27] days) and of exposure to discharge (19 [16.5-22] vs. 24 [24.5-28] days).

**Table 3.**
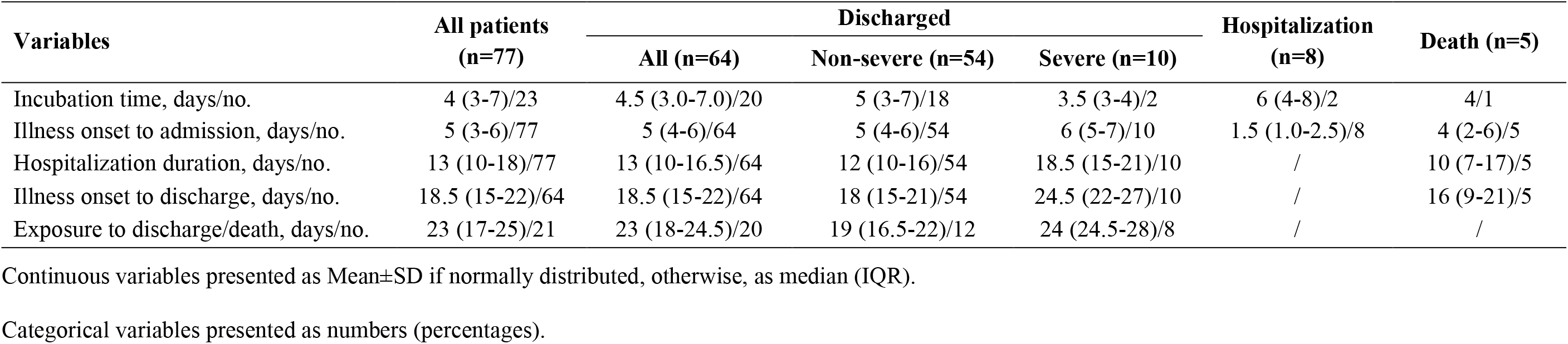
Time courses of study patients grouped by clinical outcomes.

| Variables | All patients<br>(n=77) | Discharged |  |  | Hospitalization<br>(n=8) | Death (n=5) |
| --- | --- | --- | --- | --- | --- | --- |
|  |  | All (n=64) | Non-severe (n=54) | Severe (n=10) |  |  |
| Incubation time, days/no. | 4 (3-7)/23 | 4.5 (3.0-7.0)/20 | 5 (3-7)/18 | 3.5 (3-4)/2 | 6 (4-8)/2 | 4/1 |
| Illness onset to admission, days/no. | 5 (3-6)/77 | 5 (4-6)/64 | 5 (4-6)/54 | 6 (5-7)/10 | 1.5 (1.0-2.5)/8 | 4 (2-6)/5 |
| Hospitalization duration, days/no. | 13 (10-18)/77 | 13 (10-16.5)/64 | 12 (10-16)/54 | 18.5 (15-21)/10 | / | 10 (7-17)/5 |
| Illness onset to discharge, days/no. | 18.5 (15-22)/64 | 18.5 (15-22)/64 | 18 (15-21)/54 | 24.5 (22-27)/10 | / | 16 (9-21)/5 |
| Exposure to discharge/death, days/no. | 23 (17-25)/21 | 23 (18-24.5)/20 | 19 (16.5-22)/12 | 24 (24.5-28)/8 | / | / |

Continuous variables presented as Mean±SD if normally distributed, otherwise, as median (IQR).
Categorical variables presented as numbers (percentages).

### Outcomes and risk factors for longer hospitalization of COVID-19

Follow-up were administered in all 77 patients. By the end of follow-up, 64 patients were discharged, eight remained in hospital and five died. Of five deaths, three were caused directly by COVID-19. Thirty-six patients (46.8%) were discharged within 14 days and thus reached the study endpoint, including 34 (59.6%) of non-severe patients and 2 (10%) of 20 severe patients. Overall, the cumulative probability of the endpoint was 48.3%. For non-severe and severe patients, the cumulative probabilities were 59.7% and 11.8%, respectively. Several variables with significant difference between patients with and without endpoint were selected for univariable Cox proportional-hazards regression model, as shown in Figure 2. Then all statistically significant risk factors were further included in multivariable stepwise Cox regression model, only bilateral pneumonia on CT scan, time from the illness onset to admission, severity of disease and lymphopenia remained finally, displayed in Table 4.

**Table 4.** Risk factors independently associated with the study endpoint by multivariable stepwise Cox proportional-hazards regression.

| **Risk factors** | **HR (95% CI)** | **P** |
| --- | --- | --- |
| Pneumonia on CT scan (bilateral vs. unilateral) | 0.17 (0.05-0.63) | 0.008 |
| Time from the illness onset to admission (days) | 1.45 (1.22-1.72) | <0.001 |
| Severity of disease (severe vs. non-severe) | 0.13 (0.03-0.60) | 0.009 |
| Lymphopenia (yes vs. no) | 0.17 (0.04-0.66) | 0.01 |

HR, hazards ratio; CI, confidence interval.

**Figure 2.**
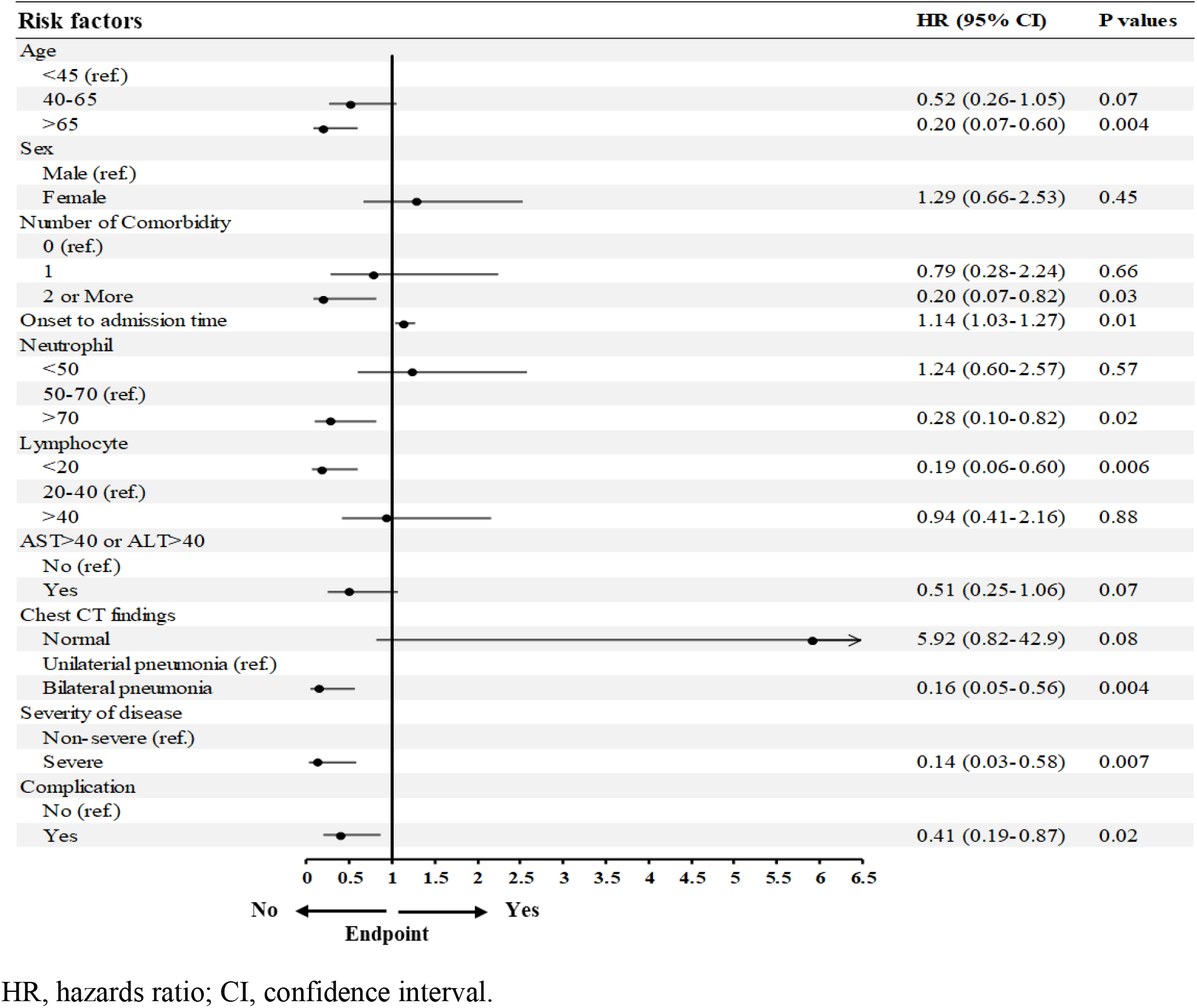
Risk factors for the study endpoint by univariable Cox proportional-hazards regression.

## Discussion

In the present study, we reported the retrospective data of 77 hospitalized patients with COVID-19 in Beijing, China, with 64 (83.1%) discharged home alive by the end of follow-up. Among a small number of patients, there was a wide span of age from 1 to 94 years, suggesting that individuals of all ages are susceptible to SARS-Cov-2 infection. Intriguingly, more females (55.8%) were identified in our study, as contract to previously studies where males were reported to constitute a higher proportion of patients ^[2–4,6]^ revealing that males had higher expression level of ACE2 receptor and thus were more susceptible to SARS-Cov-2 invasion^[14]^. Recently, Chinese Center for Disease Control and Prevention presented the biggest data of COVID-19 (44,672 confirmed cases) and indicated a relatively even sex distribution (males 51.4% vs. females 48.6%)^[15]^. These findings suggest that the sex difference found at the early stage of the outbreak might be of coincidence such as the exposure difference between sexes. In our study, most of the patients had a history of exposure to Wuhan, especially at the initial stage of the spreading in Beijing, and the number of those patients was reducing over time as along with reducing of the number of all patients. This suggested the effectiveness of quarantine of Wuhan city on the control and containment of COVID-19 in Beijing and other cities and countries as demonstrated by a more recent study^[16]^. Lots of patients (42.9%) got infected by family clustering, which emphasizes the importance of the self-isolation of confirmed and suspected cases, as well as the importance of public awareness campaign and education.

The incubation time of SARS-COV-2 has been reported by many previous studies. Li elal.^[3]^ studied the first 425 patients in Wuhan, China, and reported that the incubation period was 5.2 days with 95% confidence interval 4.1-7 days. Based on a national dataset of 1099 patients, Guan et al. ^[6]^ reported a similar period, 4 days with interquartile range 2 to 7. We obtained consistent results, the mean duration of 4 days with IQR 3-7 days. The longest incubation period in our study is 14 days, which further supports the 14-day isolation for suspected cases.

The duration of the illness onset to hospital admission is an important factor in the spreading of virus infection. The sooner patients are hospitalized and treated, the less spreading there will be. Previous studies reported that the duration of the illness onset to hospital admission in the early stage of this outbreak was 7 to 12.5 days ^[2,3,5]^ In the present study, the time from the symptom onset to hospital admission was significantly shorter (5 days, IQR 3-6 days), which should be attributed to the increased awareness of the public on the epidemic and the daily public-wide educational campaigns on precautionary measures against exposure to SARS-Cov-2. It is equivocal that whether longer duration between symptom onset to hospital admission is associated with clinical outcome of patients. Huang et al.^[2]^ did not find a difference of this duration between ICU and non-ICU patients, however, Wang et al.^[5]^ investigated more patients and revealed that patients admitted to ICU had longer duration (8 vs. 6 days, P=0.009). In the present study, we also observed a longer duration in severe patients versus non-severe patients (supplementary files). This may be attributed to that longer pre-hospital duration may delay the treatment thus lead to unfavorable outcomes.

Currently, few studies reported the average hospital length of stay of discharged patients COVID-19. In the present study, we reported the average hospital length of stay were 13 days (IQR: 10-16.5) for patients with COVID-19, which is three days longer than Wang et al.’s report ^[5]^ based on 38 discharged patients from Wuhan. However, in their study, the patients’ incubation period is two days longer than that of our study. Nevertheless, length of hospital stay of COVID-19 is significantly shorter than that of SARS in 2003, which was almost one month ^[17]^, further indicating the milder severity of COVID-19. We also revealed that the cumulative probability of COVID-19 patients discharged within two weeks were 59.7% for non-severe and 11.8% for severe patients, respectively, which means that nearly 60% of non-severe patients will be discharged within two weeks. Since length of hospital stay is related to duration of illness onset to hospital admission, duration of illness onset to discharge may be better to present the clinical course of COVID-19. If further taking incubation time into consideration, the whole duration of COVID-19, from exposure to recovery, is 23 days. To the best of our knowledge, this might be the first description of whole duration of COVID-19. But only a small number of patients from a single center were included (n=20), further studies based on large sample size from multiple centers are warranted to confirm this finding.

Longer duration of disease/hospitalization means more medical burden, especially when facing the exponentially increasing need within a short period produced by transmissible diseases like COVID-19. One of the most challenged situations during addressing the outbreak of transmissible disease might be rapidly increasing patients that excessively exceeds healthcare capacities. Although the outbreak of COVID-19 has been well controlled in China, many other countries are still facing a serious outbreak. The information reported by the present study, especially the duration of COVID-19 and the corresponding risk factors for longer hospitalization duration, could contribute to those countries to well prepare and redistribute their medical resource so as to flatten the curve and thus delay the epidemic peak.

This study has several limitations. First, a relatively small number of patients from a single center in Beijing were included. Obviously, length of hospital stay differs across hospitals, cities, and countries. It would be more valuable to draw the corresponding durations and to identify the risk factors based on the dataset at city or nation scale. Second, we could not provide useful information on the average duration from the illness onset to death, because of limited data in our study. Third, treatment is a pivotal determinant of durations of disease, however, we currently have no detailed data of treatment for patients with COVID-19 and cannot determine the influence of treatment on durations of COVID-19. But we will continue this work in the future.

In conclusion, the average hospital length of stay is 13 days for discharged patients with COVID-19; average time of clinical course is 23 days. Both are significantly shorter than SARS. Bilateral pneumonia on CT scan, shorter period of illness onset to admission, lymphopenia, severity of disease are the risk factors for longer hospitalization duration of COVID-19. There findings might be useful for the countries or territories facing the threat of COVID-19 to well prepare and rebalance their medical resource.

## Data Availability

The datasets generated during and/or analysed during the current study are available from the corresponding author on reasonable request.

## Acknowledgement

We thank all patients, researchers, doctors, and nurses involved in this study.

